# Development and validation of an end-to-end deep learning pipeline to measure pericardial effusion in echocardiography

**DOI:** 10.1101/2022.08.13.22278732

**Authors:** Cheng-Ching Wu, Chi-Yung Cheng, Huang-Chung Chen, Chun-Huei Hung, Tien-Yu Chen, Chun-Hung Richard Lin, I-Min Chiu

**Author notes:** Correspondence: I-Min Chiu. Cheng-Ching Wu and Chi-Yung Cheng contributed equally to this work.

## Abstract

**Introduction:** Cardiac tamponade, caused by pericardial effusion (PE), is a life-threatening condition that can be resolved by timely pericardiocentesis. Nevertheless, PE measurement remains operator-dependent and may be difficult in some circumstances. Our study aimed to develop a deep-learning pipeline that measures the amount of PE based on raw echocardiography clips.

**Methods:** Echocardiographic examination data were collected from one medical center in southern Taiwan from 2010–2018. Four commonly used cardiac windows, including the parasternal long-axis, parasternal short-axis, apical four-chamber, and subcostal views from included ultrasound examinations, were used for analysis. We proposed a deep learning pipeline consisting of three steps: moving window view selection, automated segmentation, and width calculation from a segmented mask. The pipeline was then prospectively validated from 2019–2020 using a dataset from the same hospital, and externally validated using data from another medical center in Taiwan. Model performance was evaluated using mean absolute error, intraclass correlation coefficient (ICC), and R-squared value between the ground truth and predictions.

**Results:** In this study, 995 echocardiographic examinations were included. Among these, 155 were used for internal validation and 258 were used for external validation. The proposed pipeline had a predictive performance of ICC=0.867 for internal validation and ICC=0.801 for external validation. It accurately detected PE with an area under the receiving operating characteristic curve (AUC) of 0.926 (0.902–0.951) for internal validation and 0.842 (0.794–0.889) for external validation. Regarding the recognition of moderate PE or worse, the AUC values improved to 0.941 (0.923–-0.960) and 0.907 (0.876–0.943) for internal and external validation, respectively. Of all the selected cardiac windows, our model had the best prediction in the parasternal long-axis and apical four-chamber views.

**Conclusions:** The machine-learning pipeline could automatically calculate the width of the PE from raw ultrasound clips. The novel concepts of moving window view selection for image quality control and computer vision techniques for maximal PE width calculation seem useful in the field of ultrasound.

## Introduction

Pericardial effusion (PE) is an acute or chronic accumulation of fluid within the pericardial space which is usually revealed by transthoracic echocardiography. Fluid accumulation increases pressure in the pericardial sac, leading to compression of the heart and subsequent cardiac tamponade. In acute settings, only 100–150 mL of fluid is necessary for cardiac tamponade to occur, which results in impaired diastolic filling and reduced cardiac output. PE is a life-threatening issue that can be resolved by timely pericardiocentesis.^1^ Therefore, the early detection of PE and measurement of PE width are important.

Echocardiography remains the gold standard imaging modality for verifying the presence of PE by demonstrating fluid collection in the pericardial space.^2,3^ It is considered the first-line imaging choice since it is less costly, more portable, more widely available, and can provide a comprehensive anatomical and functional assessment compared with computed tomography and magnetic resonance imaging.^4,5^ Nevertheless, the presence and grading of PE is associated with intraoperative uncertainty. For example, mild PE does not always correspond to true effusion and can be indicative of pericardial fat.^6^ Furthermore, the image quality of a transthoracic echocardiogram might be compromised by elements such as the female breast or obscuration by bone or lung.^4^ Early disclosure of the precise PE grade might be difficult in some circumstances and is dependent on operator experience.

Artificial intelligence (AI) has been used in many clinical settings to assist in the diagnosis of conditions based on echocardiograms. Considerable effort has been devoted to themes such as left ventricular function assessment, regional wall motion abnormality, right ventricular function, valvular heart disease, cardiomyopathy, and intracardiac mass.^7-10^ Regarding the diagnosis of PE, one study in 2020 that used a deep learning model to detect PE in echocardiography achieved an accuracy of 0.87–0.9.^11^ In daily practice, further information on PE width and severity is critical for initializing the necessary interventions. To the best of our knowledge, no study has analyzed the grading of PE via machine learning. Thus, our study attempted to develop a deep learning model using echocardiography for PE detection and PE width measurement. In addition, to better deploy the deep learning model, we propose an end-to-end guideline that can output the prediction results from raw ultrasound files.

## Method

The data collection and protocols utilized in this study were authorized by the Institutional Review Board of E-Da Hospital (EDH; no: EMRP24110N) and the Institutional Review Board of Kaohsiung Chang Gung Memorial Hospital (CGMH; no: 20211889B0 and 202101662B0).

### Data Collection

In this study, images from routine echocardiography were generated at two medical centers, EDH and CGMH, in southern Taiwan. The deep learning model was trained and internally validated in EDH and externally validated in CGMH.

During data collection, we used “pericardial effusion” as a keyword to search the Hybrid picturE Report System in EDH to collect the examination list. We obtained patients’ raw data from transthoracic echocardiography examinations with PE performed at EDH between January 1, 2010, and June 30, 2020. These data were divided into training and validation datasets based on the respective index dates of the examinations. Examinations with index dates prior to December 31, 2018, were used for the development of the model, and examinations with index dates after January 1, 2019, were used for internal validation. To test the generalizability of the model, we retrieved echocardiography data from CGMH between January 1, 2019, and June 30, 2020, for external validation. The study flowchart and data summary are presented in Figure 1.

**Figure 1.**
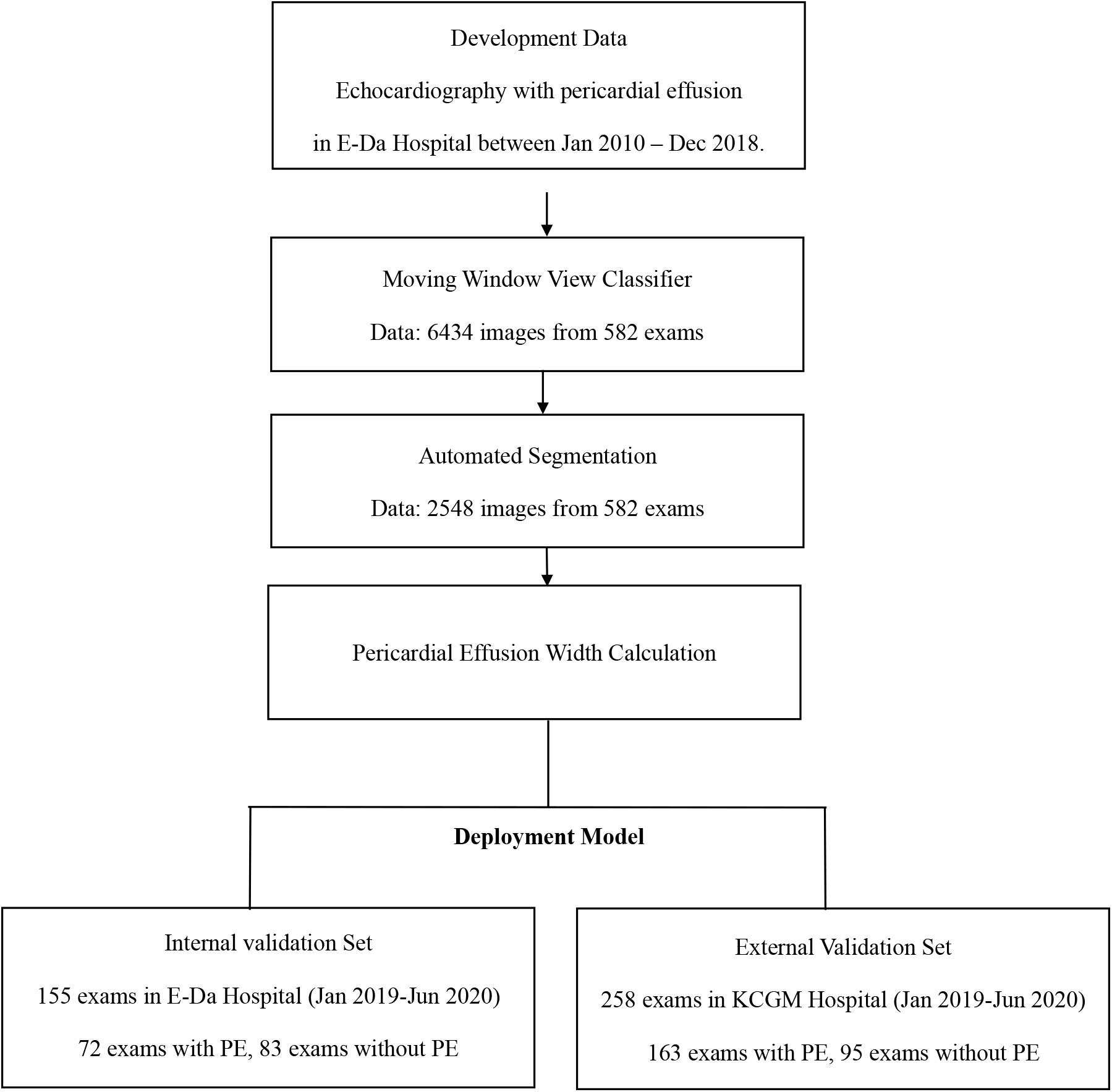
Flow chart illustrating the study design and data summary.

### Echocardiography

Images were gathered in a normal manner, with patients lying in the left lateral decubitus position. The ultrasound system (IE33, Philips Healthcare; S70, GE Healthcare; or SC2000, Siemens Healthineers) was used to perform echocardiographic examinations in EDH. Data from CGMH for external validation were acquired using EPIC7 (Philips Healthcare), Vivid E9 (GE Healthcare), or SC2000 (Siemens Healthineers). All examinations were saved in picture archiving and communication systems in the Digital Imaging and Communications in Medicine (DICOM) format.

After extracting the raw DICOM files, we processed the image from each patient to select the proper echocardiography views for developing a deep learning pipeline. The selected views were the parasternal long-axis (PLAX), parasternal short-axis (PSAX), apical four-chamber (A4C), and subcostal (SC) views. The ground truth of PE width was annotated from the examination report by a proficient cardiac physiologist and was inspected by a cardiologist who delegated the confirmed reports.

### Deep Learning Model Development

In this study, we developed an end-to-end pipeline for the automated measurement of PE based on the steps outlined below (Figure 2). The training subset of videos from EDH was used for the three main tasks of our pipeline:

**Figure 2.**
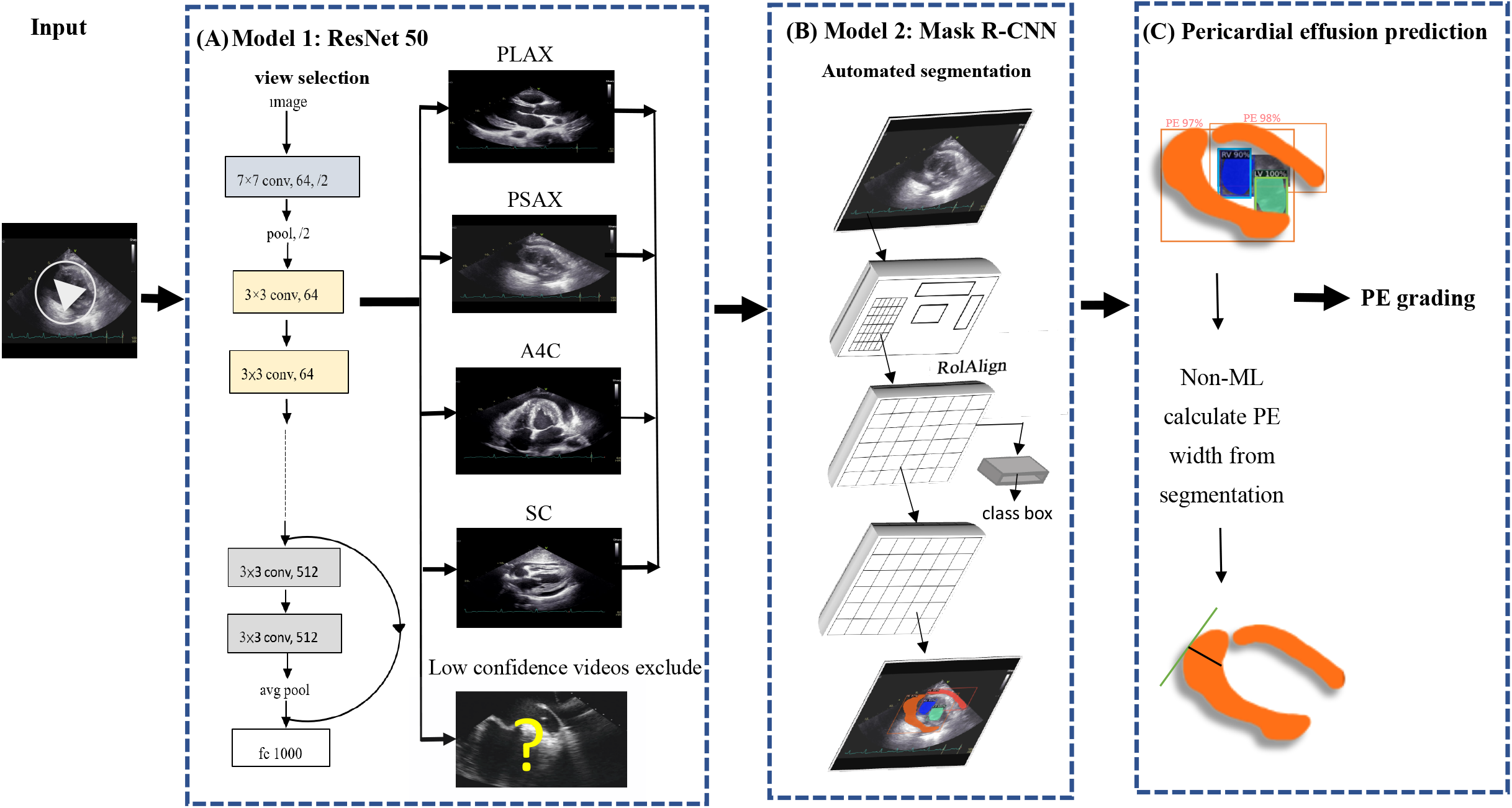
Deep Learning Pipeline for PE Measurement. This graphic demonstrated the end-to-end pipeline proposed in this study. The pipeline consisted of three major steps, which were moving window view selection (Figure 2A), automated PE segmentation (Figure 2B), and PE width calculation from segmented mask (Figure 3B). The pipeline was able to process raw DICOM file from echocardiography exam directly and output PE width prediction.

### Step one: moving window view selection (MWVS)

We proposed a pipeline to directly manage echocardiography files from the workstation, similar to the work done by Zhang et al. and Huang et al., with some adjustments (Figure 2A).^10,12^ To distinguish the four primary views (PLAX, PSAX, A4C, and SC) from other views during each examination, we developed the first deep neural network model. This model was a ResNet-50-based two-dimensional model that aimed to classify each frame from the extracted DICOM files of echocardiography into the selected view types.^13^ To train this model, we randomly selected 6,434 images from the training dataset of EDH and labeled them according to the four primary views or other views, including low-quality views. We trained the model with data splitting of 80% and 20% for the training and validation sets, respectively. The model weight with the best prediction performance in the validation set during the training process was preserved. The prediction accuracy was assessed for each view class and weighted average result.

While managing the input video from the patient, we used 48 frames moving window to filter all videos. For each video, we retrieved a clip of 48 frames with the best confidence with regard to the specific view type by majority voting (Figure 3). The MWVS concept was used not only as a view classifier but also for quality control. This process not only helps the algorithm to identify the right video but also retrieves the best 48 frames of the video with regard to image quality. If a video did not contain any of the 48 consecutive frames that consisted of qualified frames higher than 50% from one of the four primary views, it was excluded from further analysis. Moreover, the view-classifying confidence levels for all images obtained from the clip of 48 frames were averaged to evaluate the general image quality, and they correlated with further performance. Ultrasound videos with an average confidence level of < 0.8 were excluded from further automated segmentation.

**Figure 3.**
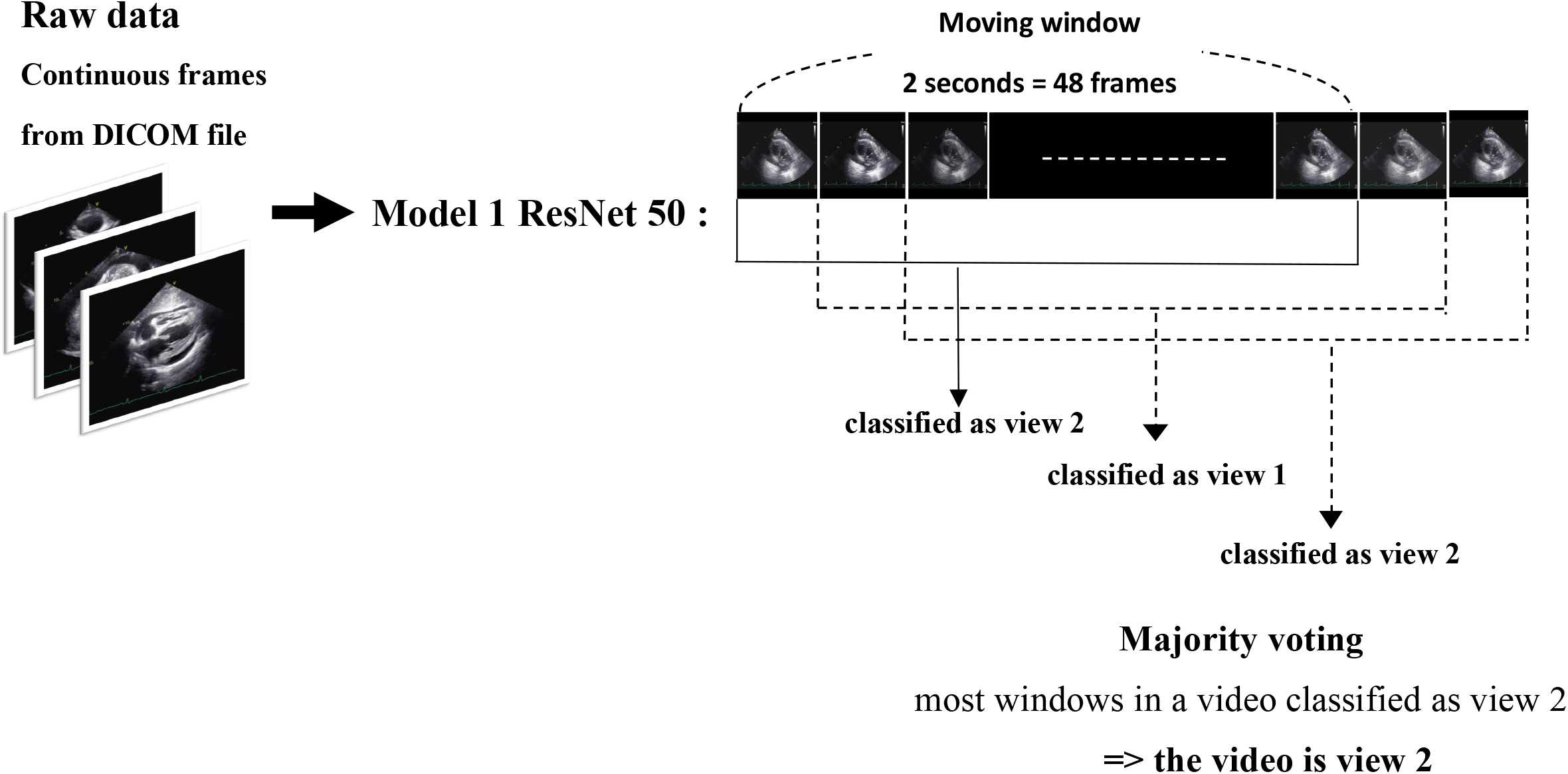
Moving Window View Selection. We used 48 continuous frames as window to iterate through raw video. In each window, the view will be classified by majority voting from the prediction of all48 frames. For example, if most frames in the window were classified as view 2, the window was classified as view 2. As the window iterate through raw video, the window with highest confidence of classification will be preserved for further analysis.

### Step two: automated segmentation

From the dataset, we annotated 2,548 randomly selected frames in the EDH training dataset which were evenly distributed over the four primary views. For each view, we manually labeled the segmented area for PE at three different phases in the cardiac cycle: end-systolic phase, end-diastolic phase, and middle phase between the two aforementioned phases. We also labeled the segmented areas for the four cardiac chambers to enhance the model performance in separating these fluid-containing areas. We used a mask region-convolutional neural network (R-CNN) as the framework to train object instance segmentation based on the labeled ground truth (Figure 2B). The model was trained with 80% and 20% data splitting for the training and validation sets, respectively. Mask R-CNN is commonly used for instance segmentation tasks in medical applications because it can simultaneously perform pixel-level segmentation and classification of multiple target lesions.^14^ The implemented model generates bounding boxes and targeting masks for each instance of an object in an image. As such, the input comprised consecutive ultrasound frames, and the output comprised a segmented mask, which indicated the corresponding four cardiac chambers and PE. The accuracy of the segmentation model was assessed using the Dice coefficient metric.

### Step three: measurement of pericardial effusion

After generating a segmented mask for PE, we proposed a computer vision technique (maximal width calculator of the segmented mask) to calculate the largest width of the PE in each ultrasound frame (Figure 2C). For this task, we iterated through the vertical axis in each frame and hypothetically drew a horizontal line to see if there was any intersection between the segmented mask and horizontal line. If an intersection existed, we obtained a normal line from the edge of the mask over the intersection point. The length of the normal line that passes through the segmented mask was counted as the width of the PE at that intersection point. The largest width of the PE through the iteration over the vertical axis was regarded as the width of the PE of the frame. The same technique was applied to all 48 frames in the ultrasound video to provide the optimal PE width.

### Statistical Analysis

Continuous variables are presented as mean (standard deviation) if normally distributed, otherwise they are presented as median with the interquartile range. Dichotomous data are presented as numbers (percentage). Categorical variables were analyzed using the χ2 test. Continuous variables were analyzed using the independent-sample t-test if normally distributed; otherwise, the Mann-Whitney U test was used.

Model performance regarding PE width measurement was analyzed based on the mean absolute error, intraclass correlation coefficient (ICC), and R-square value between the ground truth and prediction. We further examined the prediction of the existence of PE and moderate PE using sensitivity, specificity, and area under the receiver operating characteristic curve (AUC). The deep learning models in the proposed pipeline were developed using the TensorFlow Python package. Image manipulation was performed using OpenCV 3.0 and scikit images. All analyses were performed using SPSS for MAC version 26.

## Results

In this study, 737 examinations from EDH were included in the analysis, of which 582 were included in the training set and 155 were in the internal validation set. For the external validation, 258 examinations from CGMH were included. Because there were less than 10 SC views in the CGMH dataset, the SC view was excluded from further analysis in the external validation.

The demographic and clinical characteristics of those who underwent echocardiography are presented in Table 1. The mean ages of patients in the training, internal validation, and external validation set were 67.4±15.4, 59.8±19.2, and 66.4±16.1 years, respectively; 46.5% and 63.2% of patients in the internal and external validation groups, respectively, had PE. The average ejection fraction was 64.3±7.1% in the internal validation group and 61±13.9% in external validation group.

**Table 1.**
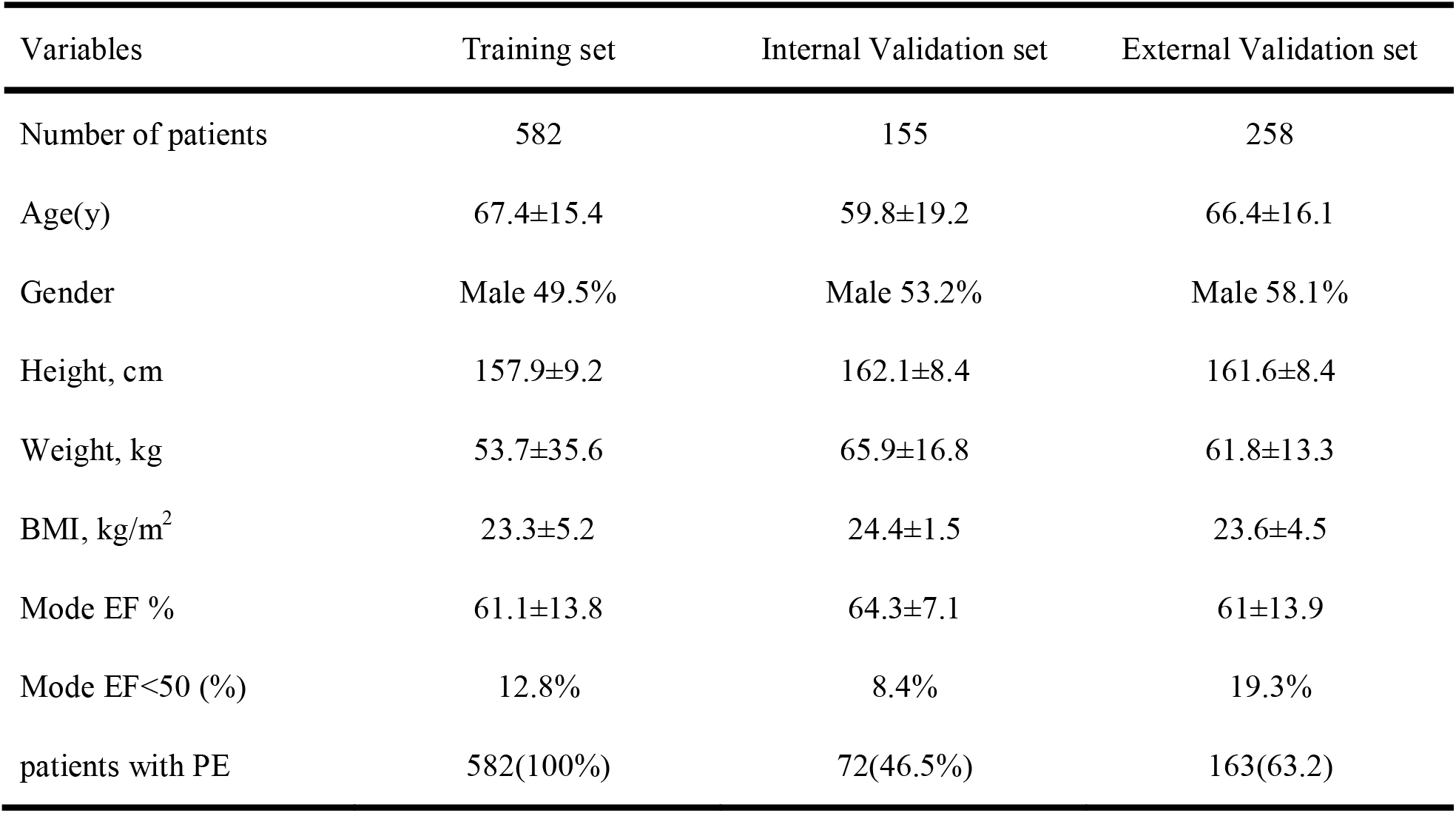
Demographics, Basic Characteristics, and Clinical Findings of the patients

| Variables | Training set | Internal Validation set | External Validation set |
| --- | --- | --- | --- |
| Number of patients | 582 | 155 | 258 |
| Age(y) | 67.4±15.4 | 59.8±19.2 | 66.4±16.1 |
| Gender | Male 49.5% | Male 53.2% | Male 58.1% |
| Height, cm | 157.9±9.2 | 162.1±8.4 | 161.6±8.4 |
| Weight, kg | 53.7±35.6 | 65.9±16.8 | 61.8±13.3 |
| BMI, kg/m<sup>2</sup> | 23.3±5.2 | 24.4±1.5 | 23.6±4.5 |
| Mode EF % | 61.1±13.8 | 64.3±7.1 | 61±13.9 |
| Mode EF<50 (%) | 12.8% | 8.4% | 19.3% |
| patients with PE | 582(100%) | 72(46.5%) | 163(63.2) |

The view classifier achieved an average accuracy of 0.91 and 0.87 in predicting image classes in the training and validation sets, respectively. The independent accuracies in the validation set for each class were 0.90, 0.87, 0.93, 0.76, and 0.88 for PLAX, PSAX, A4C, SC, and others, respectively.

After MWVS, most of the ultrasound videos, ranging from 80–100% among the four selected ultrasound views in EDH, successfully passed through for the segmentation model. In the external validation (CGMH dataset), 686 ultrasound videos from the four selected views were obtained. Our MWVS scanned through all DICOM files of 258 patients, and 53.2% of the 686 ultrasound videos were preserved for segmentation inference. The videos selected by our pipeline were further checked by a cardiologist, and none of them were misclassified into other cardiac views.

In image segmentation, the mask R-CNN-based model effectively localized the cardiac chambers and PE area within the four different views (Figure 4). In the validation set consisting of 510 images, the average Dice coefficient ranged from 0.67–0.82 among the four different views, with the SC view being the lowest. PE segmentation in the PLAX view showed the best Dice result 0.72, while the SC view had the poorest result (0.56) (Table 2). Based on the segmentation of the PE area, we calculated the maximal PE width from each frame.

**Figure 4.**
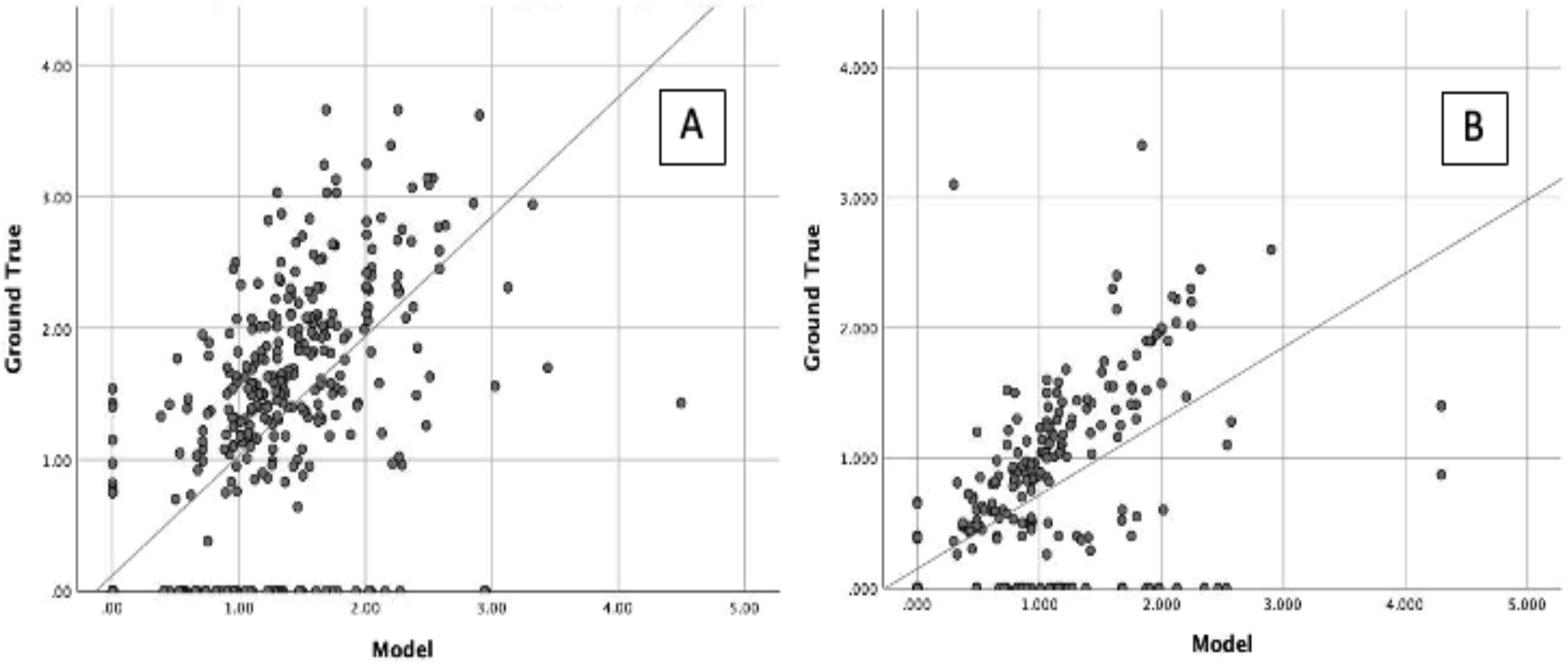
Scatter plot of PE prediction from deep learning model compare with human expert. Figure 4A represent the scatter plot from internal validation, and Figure 4B from external validation.

**Table 2.** Dice coefficient of image segmentation

| Dice Coefficient |  |  |  |  |  |  |
| --- | --- | --- | --- | --- | --- | --- |
|  | PE | RV | LV | RA | LA | Average |
| PLAX | 0.72 | 0.86 | 0.85 |  | 0.84 | 0.82 |
| PSAX | 0.69 | 0.59 | 0.85 |  |  | 0.71 |
| A4C | 0.58 | 0.81 | 0.86 | 0.82 | 0.83 | 0.78 |
| SC | 0.56 | 0.66 | 0.71 | 0.70 | 0.72 | 0.67 |

Figure 4 shows the scatter plot of the PE width measurement between the ground truth and model prediction after finding the largest normal line passing the segmented mask in each frame. Compared with the ground truth, we reported the absolute difference and correlation between automated and manual measurements of PE in both the internal (EDH) and external (CGMH) validation datasets. The mean absolute error was 0.33 cm and 0.35 cm in internal and external datasets, respectively. Interobserver variability was highly correlated for the measurement of PE width between our model and human expert (ICC=0.867, p<0.001, EDH; ICC=0.801, p<0.001, CGMH). The R2 was 0.594 for EDH and 0.488 for CGMH validation dataset.

Our model accurately detected the existence of PE in the internal validation (AUC=0.926 [0.902–0.951]) and external validation (AUC=0.842 [0.794–0.889]). With regard to recognizing moderate PE or worse, the AUC values improved to 0.941 (0.923–-0.960) and 0.907 (0.876–0.943) in the internal and external validation groups, respectively.

We further performed a stratified analysis of the model prediction in the different echocardiography views. In the internal validation, the model prediction of PE width was highly correlated with the ground truth in the four different views, with ICC ranging from 0.802–0.910. The PLAX and A4C views appeared to have the best prediction results with ICCs of 0.910 (0.876–0.935) and 0.907 (0.871–0.932), respectively. In the external validation, similar to internal validation, the model performed better in the PLAX and A4C views, with ICCs of 0.807 (0.726–0.864) and 0.897 (0.846–0.931), respectively. The other performances are listed in Table 3.

**Table 3.**

|  | number before view<br>selection | number after moving<br>window selection | Mean Absolute Error<br>(cm) | ICC | R2 |
| --- | --- | --- | --- | --- | --- |
| **Internal validation** |  |  |  |  |  |
| PLAX | n=155 | n=146 | 0.28 | 0.910 (0.876-0.935) | 0.700 |
| PSAX | n=155 | n=155 | 0.46 | 0.802 (0.728-0.856) | 0.469 |
| A4C | n=155 | n=155 | 0.32 | 0.907 (0.871-0.932) | 0.754 |
| SC | n=155 | n=124 | 0.40 | 0.865 (0.808-0.905) | 0.590 |
| **External validation** |  |  |  |  |  |
| PLAX | 222 | n=127 | 0.32 | 0.807 (0.726-0.864) | 0.457 |
| PSAX | 252 | n=138 | 0.44 | 0.714 (0.600-0.796) | 0.337 |
| A4C | 212 | n=100 | 0.11 | 0.897 (0.846-0.931) | 0.662 |

## Discussion

Computer vision and deep learning models have proven useful for aiding echocardiography interpretation, estimating cardiac function, and identifying local cardiac structures. In recent years, deep learning algorithms have also been applied to facilitate the diagnosis of PE.^15^ Nayak et al. developed a CNN that detected PE in the A4C and SC views with accuracies of 91% and 87%, respectively.^11^ Pericardiocentesis is an essential therapeutic procedure for the treatment of symptomatic PE. However, this therapeutic procedure can be life-threatening. The major complications of pericardiocentesis are mortality, cardiac arrest, cardiac perforation, and cardiac chamber laceration, while other complications include pneumothorax, supraventricular tachycardia, and pneumopericardial fistula.^16^ Blind pericardiocentesis is associated with a mortality rate of 6% and complication rates of 20–50%^17,18^. With ultrasound-assisted pericardiocentesis, the mortality rate is <1%, and the overall complication rate is approximately 4–20%.^18-20^ Therefore, it is important to identify the location and distribution of pericardial fluid while avoiding the accidental puncture of vital organs.

In this study, we proposed a machine learning pipeline that could process raw DICOM files from ultrasound and predict the PE width in clinical practice. This pipeline combines two steps of the deep learning model and one technical calculation algorithm to accurately predict PE width. Few efforts have been made to predict PE existence,^11^ with some studies being based on computed tomography scans.^21,22^ To the best of our knowledge, this is the first video-based machine learning model to predict the PE width using echocardiography. The correlation between the prediction of our model and human experts was highly in both the internal and external validation datasets, with the best performance noted in the PLAX view. The speed of inference from accessing the file to the output for our model in one graphics processing unit (GPU; NVIDIA RTX 3090) was approximately 30–40 s for one examination, which is usually faster than human assessment.

The two methods proposed in this study are novel concepts, including MWVS for view selection and the maximal width calculator of the segmented mask. These two methods are crucial for real-world predictions, particularly for relatively smaller datasets. Many previous studies used datasets manually selected by human experts during dataset cleaning for machine learning and used only “textbook-quality” images for training.^23-26^ In contrast, we hypothesized that an analytic pipeline could automatically analyze echocardiograms and be easily applied to personal devices or web applications. Hence, it is important to exclude processes that require an expert sonographer or a cardiologist. Madani et al. trained a CNN to simultaneously classify 15 standard echocardiogram views acquired based on a range of real-world clinical variations, and the model showed high accuracy for view classification.^19^ Similarly, in our study, we used echocardiogram video clips randomly obtained from the real world, which were taken for a variety of clinical purposes, including ejection fraction calculation and for detecting PE, valve disease, regional wall abnormality, cardiomyopathy, and pulmonary arterial hypertension. We developed an initial screening model for view classification and quality control. All raw images from the medical image database were input into the screening model, leaving a specific view of sufficient quality for diagnosis. In addition, with the “moving window” concept, we retrieved only clips with 48 consecutive frames that fulfilled the image quality. We found that ICC and diagnostic accuracy were significantly improved after MWVS. By avoiding limited or idealized training datasets, we believe that this model is broadly applicable to clinical practice.

Rather than previous studies using three-dimensional CNN architecture for training the view classifier,^10^ we used a two-dimensional ResNet structure. The two-dimensional structure consumes significantly less computing resources and can be deployed as a real-time feedback system for ultrasound operators using only one GPU. We proposed MWVS by combining a two-dimensional image classifier with a moving-window algorithm for clinical usage. MWVS is a novel concept that has not yet been proposed in the field of echocardiography assisted by machine learning. MWVS plays the role of an image quality filter, and the major function of MWVS is to ensure image quality in keeping with the next step in the pipeline. In EDH, echocardiography is performed by well-trained technicians who follow the protocol designed by the echocardiologist consensus committee. Therefore, the original images from EDH had good homogeneity, and MWVS filtered out fewer patients. At CGMH, echocardiography is performed by a separate echocardiologist who has the respective habit to perform echo study. Therefore, the original images from CGMH had poor homogeneity, and MWVS filtered out more patients. This finding proved that MWVS plays a significant role in maintaining image quality. However, this finding also confirms that the applicability of machine learning depends on image homogeneity.

After segmenting the PE, we developed a computer vision technique to calculate the largest PE width. The current categorization of PE size depends on linear measurements of the largest width of the effusion at end-diastole and is graded as small (<1 cm), moderate (1–2 cm), and large (>2 cm).^27^ This semiquantitative classification method is prone to errors because of the asymmetric loculated effusion and shifts in fluid location during the cardiac cycle.^28^ Therefore, an automated calculation system could help identify the largest width of the PE in every ultrasound frame without any errors. Compared with AI-based models, the computer vision technique is more similar to the method used by human experts. The AI-based model not only consumes more computing resources but also requires a large number of datasets for training and validation. To our knowledge, our study is the first to use computer vision techniques for classifying PE grades.

This study had certain limitations. First, we conducted the study retrospectively and trained our model using only one hospital dataset. The sample size and ethnic diversity were limited and the results may not be generalizable to other areas. The dataset should have greater heterogeneity in a multicenter setting. However, the enrolled images were obtained using different ultrasonography machines operated by several echocardiographers. Moreover, we used standard views for diagnosis, and model achievement was similar during external validation. Second, although we graded the amount of PE, there was no information on whether there was a sign of cardiac tamponade, because PE volume does not necessarily correlate with clinical symptoms.^29^ Further research should also evaluate the collapsibility of the cardiac chambers and the presence of tamponade signs.

## Conclusion

We developed a machine-learning pipeline that automatically calculates the width of the PE from raw ultrasound clips. The model achieved high accuracy in detecting PE and predicting the PE width in both internal and external validation. The concept of MWVS for image quality and computer vision techniques for maximal PE width calculators is a novel application in the field of ultrasound.

## Data Availability

Data sharing is not applicable to this article.

## Abbreviations

A4C: apical four-chamber
AI: artificial intelligence
AUC: area under the receiver operating characteristics curve
CGMH: Chang Gung Memorial Hospital
CNN: convolutional neural network
DICOM: Digital Imaging and Communications in Medicine
EDH: E-Da Hospital
GPU: graphics processing unit
ICC: intraclass correlation coefficient
MWVS: moving window view selection
PE: pleural effusion
PLAX: parasternal long-axis
PSAX: parasternal short-axis
SC: subcosta

## Acknowledgements

We appreciated the Biostatistics Center, Kaohsiung Chang Gung Memorial Hospital for statistics work.

